# Genetically Predicted Inflammatory Proteins Mediate the Association Between Gut Microbiota and Preterm Delivery: A Mendelian Randomization Study

**DOI:** 10.1101/2024.10.22.24315951

**Authors:** Min Zhang, Xiaodan Chen, Yan Zhang, Jia Huang, Ling Chen

**Author notes:** **Correspondence:** Min Zhang. Department of Preventive Healthcare, Datuan Community Health Service Center. 169 Yongding South Street, Pudong New District, Shanghai 201311, China. **Xiaodan Chen:**. **Yan Zhang:**. **Jia Huang:**. **Ling Chen:**.

## Abstract

**Aims:** Inflammatory proteins and unique gut microbiota profiles characterize preterm delivery (PTD). Nevertheless, the comprehensive understanding of gut microbiota and inflammatory proteins of PTD remains unclear. This study aimed to investigate the causal relationship between gut microbiota and PTD and identify the inflammatory proteins as potential mediators.

**Methods and results:** The exposure genome-wide association studies (GWAS) data were sourced from the GWAS Catalog, while the outcome GWAS data were obtained from the Early Growth Genetics (EGG) Consortium. The study used 473 types of gut microbiota, 91 types of inflammatory proteins, and PTD from GWAS. We then performed two-sample Mendelian randomization (TSMR) and bidirectional Mendelian randomization (BDMR) analyses to explore the causal relationships between gut microbiota, inflammatory proteins, and PTD. Additionally, we conducted two-step Mendelian randomization (2SMR) to identify potential mediating inflammatory proteins in this process. MR analysis identified 26 gut microbiota and 6 types of inflammatory proteins causally associated with PTD. Furthermore, there was no strong evidence that genetically predicted PTD affected these gut microbiota and inflammatory proteins. Further, 2SMR analysis revealed that the association between Elusimicrobiaceae and PTD was mediated by the C-C motif chemokine 23 (CCL23), accounting for 5.09% (95%CI; 4.1%-8.7%) of the association. Similarly, the relationship between Thioalkalivibrionaceae and PTD was mediated by the Interleukin-20 receptor subunit alpha (IL-20RA), which accounted for 16.88% (95%CI; 12.77%-20.99%) of the association.

**Conclusions:** Our results reveal that Elusimicrobiaceae and Thioalkalivibrionaceae were significantly associated with PTD, with mediation occurring via CCL23 and IL-20RA, respectively.

**Impact Statement:** This study establishes a causal link between specific gut microbiota, inflammatory proteins, and PTD through MR analyses. The findings indicate that targeting the pathways involving Elusimicrobiaceae - CCL23 - PTD and Thioalkalivibrionaceae - IL20RA - PTD may provide promising interventions for preventing and treating PTD.

## 1. Introduction

Preterm delivery (PTD) refers to the delivery of a baby before 37 weeks of gestation, which can have significant implications for the health of newborns, increasing their risk of various complications, including respiratory distress, infections, feeding difficulties, and developmental disorders. According to the World Health Organization, approximately 15 million infants are born preterm each year globally.^1^ The rates of PTD vary across different countries and regions, with some low-income countries experiencing particularly high rates.^2^ The economic burden of PTD on families and society is substantial, as preterm infants often require extended medical care, which can place significant financial strain on families.^3^

The gut microbiota influences maternal nutrient absorption and metabolism, which are crucial for fetal development.^4^ An imbalance in gut microbiota may impair the absorption of essential nutrients, adversely affecting fetal growth and increasing the risk of PTD. Mounting evidence suggests that the delicate balance of the gut microbial community plays a pivotal role in the complex interplay between inflammation and the risk of PTD. Dysbiosis, characterized by a reduction in microbial diversity and an imbalance in microbial composition, has been associated with the onset of systemic low-grade inflammation.^5^ Inflammatory mediators produced by the dysbiotic gut microbiota can disrupt the maternal-fetal interface, potentially triggering preterm labor. Lipopolysaccharides (LPS), a potent pro-inflammatory molecule derived from the cell walls of certain gut bacteria, can enter the circulatory system and elicit a robust immune response, contributing to the development of PTD.^6-7^

In conclusion, the gut microbiota, inflammatory mediators, and PTD are intricately linked. ^8^ Understanding the complex mechanisms by which gut dysbiosis, inflammatory pathways, and metabolic alterations interact to influence pregnancy outcomes can provide valuable insights into potential therapeutic strategies aimed at modulating the microbiome and mitigating the risk of PTD. Nevertheless, the comprehensive understanding of gut microbiota and inflammatory proteins of PTD remains unclear.

Mendelian randomization (MR) analysis is a robust epidemiological research strategy based on Mendelian inheritance.^9-10^ This technique allows for estimating causal relationships using genetic variants as instrumental variables (IVs), thereby addressing confounding variables, measurement errors, and reverse causation.^11^ We applied three large-scale genome-wide association studies (GWAS) to study the relationship between 473 gut microbiota and 91 inflammatory proteins and PTD using MR. The aim is to elucidate the causal relationship between gut microbiota, inflammatory proteins, and PTD and to gain a clearer understanding of how gut microbiota affects PTD through its effects on inflammatory proteins.

## 2. Materials and Methods

### 2.1 Study design

In this study, we employed a range of Mendelian Randomization (MR) methodologies, including two-sample MR (TSMR), bidirectional MR (BDMR), and two-step MR (2SMR), to investigate the causal relationships between gut microbiota, inflammatory proteins, and PTD. By employing BDMR, we avoid the potential for reverse causation, ensuring that our findings accurately reflect the directionality of the relationships.^12^

Our study design is based on three key assumptions essential for the causal interpretation of MR estimates.^13^ We ensured the reliability of our findings by using genetic variations or single nucleotide polymorphisms (SNPs) as IVs that meet three crucial criteria: (i) the genetic IVs are strongly associated with exposure, (ii) the genetic IVs are not associated with confounders linked to the selected exposure and outcome, and (iii) the genetic IVs influence the outcome only through the exposure.^14^ Additionally, we completed the STROBE-MR checklist to ensure the integrity of this observational MR study;^15-16^ The relevant details are presented in Supplementary Table S1, and a clear overview of our study design is presented in Figure 1.

**Figure 1.**
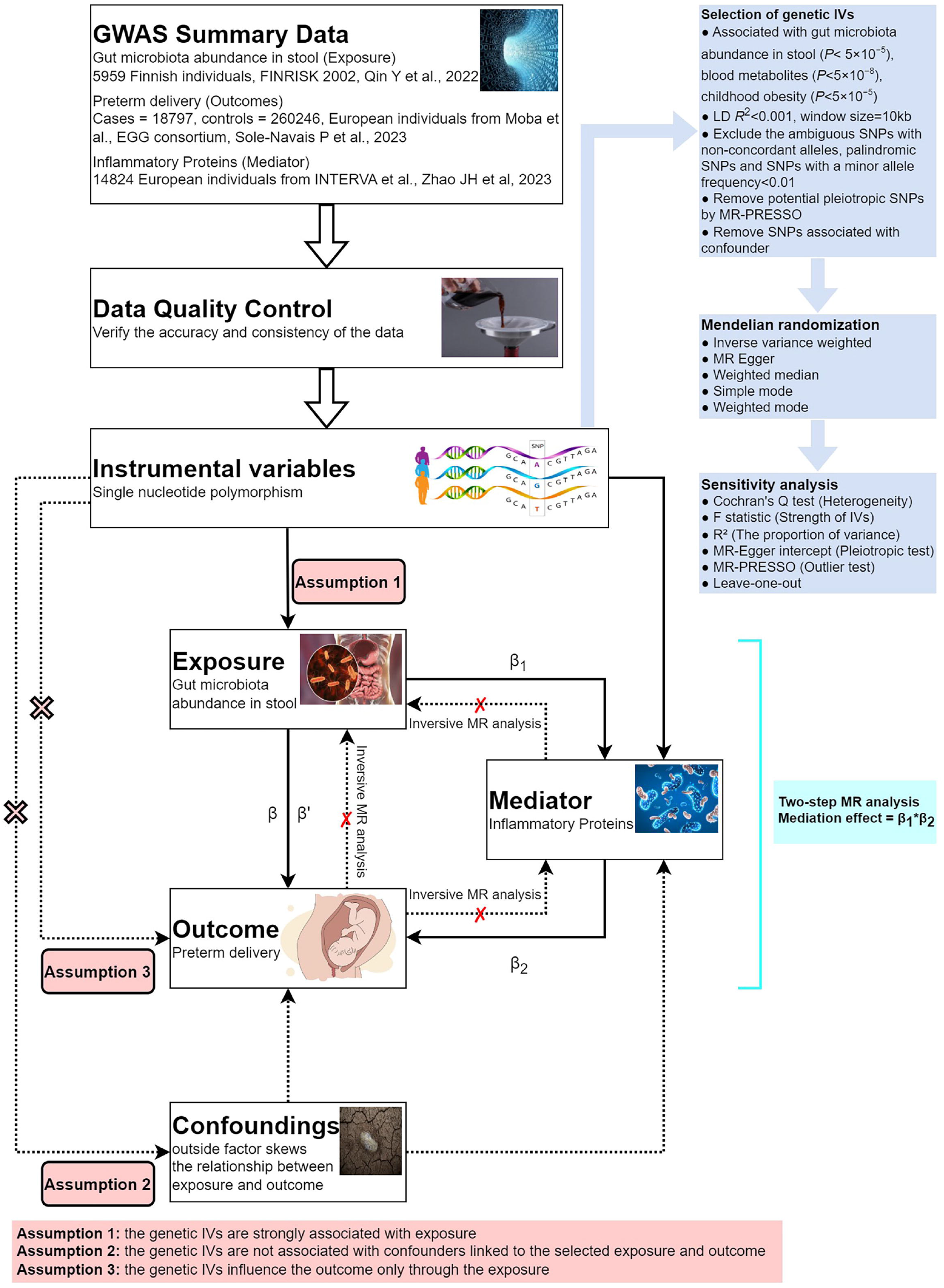
Overall study design plan. GWAS, genome-wide association studies; SNPs: single-nucleotide polymorphisms; β: the total causal effects of exposure on outcome was calculated using 2SMR method; β1: the causal effects of exposure on mediators were calculated using 2SMR method. β2: the causal effects of mediators on outcome were calculated using MVMR method. β’: the direct causal effects of exposure on outcome were calculated using the formula β’= β-(β1xβ2).

### 2.2 Data sources

Gut microbiota association studies (GWAS) data utilized in this research were obtained from a study of the genetic characteristics of gut microbiota. The original GWAS was conducted on 2,801 microbial taxa and 7,967,866 human genetic variants from 5,959 individuals enrolled in the FR02 cohort. GWAS summary data for gut microbiota was downloaded from the GWAS Catalog (https://www.ebi.ac.uk/gwas/), and the GWAS Catalog accession numbers range from GCST90032172 to GCST90032644. More detailed information about the GWAS data can be obtained from their study.^17^

The GWAS data for inflammatory proteins were sourced from the GWAS Catalog, originally generated by Zhao JH et al.,^18^ with GWAS Catalog accession numbers ranging from GCST90274758 to GCST GCST90274848. This comprehensive study identified genetic associations for 91 circulating inflammatory proteins across 11 cohorts, encompassing a total of 14,824 participants, predominantly of European ancestry.

Data on PTD has been contributed by the EGG Consortium and has been downloaded from www.egg-consortium.org.^19^ PTD was defined as a spontaneous delivery <259 days (37 completed gestational weeks) or by using the ICD-10 O60 code, and controls as a delivery occurring between 273 and 294 days (39 and 42 gestational weeks). Maternal GWAS meta-analysis of PTD in 233,290 women (n cases=15,419), all of whom are of recent European ancestry. Data from 23andMe is not included in the meta-analysis.

### 2.3 Selection of IVs and data harmonization

To adhere to the stringent criteria based on the three principal MR assumptions and to mitigate horizontal pleiotropy, only independent genome-wide significant SNPs were employed as IVs for the exposure. The IVs must be closely related to the exposure (gut microbiota, inflammatory proteins, PTD), and SNPs significantly associated with the occurrence were selected at the whole-genome level (P<5×10^−8^, r^2^<0.001, window size=10kb), If there are too few SNPs Included, the inclusion criteria can be changed to (P<5×10^−5^ or P<1×10^−6^, r^2^<0.001, window size=10kb). Additionally, we calculated the F statistics of the IVs to assess the extent of weak instrument bias. To reduce the bias caused by weak working variables, the working variables with F > 10 are retained. The formula for calculating the F value and R^2^ is as follows.

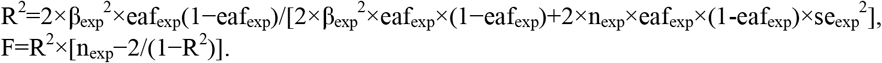

Finally, after data filtering, horizontal pleiotropy analysis was conducted. If there is horizontal pleiotropy, the MR-PRESSO test is performed.^20^ The outliers obtained from the test were removed before proceeding with further analysis.

### 2.4 Mediation analyses link “gut microbiota–inflammatory proteins–PTD”

Firstly, We performed BDMR analyses to investigate the causal relationship between gut microbiota and PTD, estimating the total effect (β) of this relationship. We used the inverse variance weighted (IVW) method to estimate effects, reporting β±SE for continuous outcomes and OR (95%CI) for binary outcomes. In brief, the IVW method meta-analyzed SNP-specific Wald estimates by dividing the SNP-outcome association by the SNP-exposure association, using random effects to derive the final causal effect estimate. Additionally, we used MR-Egger and weighted median methods as complementary approaches to IVW, providing a more comprehensive understanding of the causal relationship.

Then, we proceeded to investigate the causal relationship between inflammatory proteins and PTD using the same BDMR approach. We employed the inverse variance weighted (IVW) method, along with MR-Egger and weighted median methods, to ensure a robust understanding of this relationship.

Finally, we conducted a mediation analysis using a 2SMR study to explore whether inflammatory proteins mediate the causal effect from the gut microbiota to PTD. ^21^ The overall effect was decomposed into a direct effect (without mediators) and an indirect effect (through mediators). The total effect of gut microbiota on PTD was decomposed indirect effects of gut microbiota on PTD (overall effect) and indirect effects mediated by inflammatory proteins through the mediator (mediation effect). Mediation effects were calculated through β1×β2: (i) the causal effect of the mediator (91 inflammatory proteins) on PTD (β2) and (ii) the causal effect of the exposure (2 significant gut microbiota on PTD in primary MR analysis) on the mediator (β1). We calculated the percentage mediated by the mediating effect by dividing the indirect effect by the total effect. Meanwhile, 95% CIs were calculated with the delta method as previously reported.^22-23^

### 2.5 Sensitivity analysis

The directional association between each identified SNP and both the exposure and outcome variables was assessed using MR Steiger filtering.^24^ This method measures the degree to which the variation in exposure and outcomes can be attributed to instrumental SNPs and determines if the variability in outcomes is less than that in exposure. Horizontal pleiotropy was further investigated via the MR-Egger approach, which utilizes weighted linear regression with an unconstrained intercept. ^25^ This intercept acts as an indicator of the average pleiotropic effect across genetic variations, reflecting the typical direct influence of a variant on the outcome variable. If the intercept significantly deviated from zero (MR-Egger intercept p<0.05), it indicated the presence of horizontal pleiotropy. Moreover, Cochrane’s Q-test was used to assess heterogeneity, with lower p-values suggesting increased heterogeneity and a higher probability of directional pleiotropy. Leave-one-out analyses were also performed to identify potential SNP outliers.^26^

### 2.6 Statistical analysis

We used the results calculated by the IVW method as the final main result, and a significance threshold of p<0.05 was applied to the MR analysis, where P-values below this threshold were considered statistically significant. All statistical analyses and data visualizations were performed using R software (R Foundation, Vienna, Austria), with the TwoSampleMR (https://github.com/MRCIEU/TwoSampleMR) package for 2SMR analysis and the PNG (Boutell, Netherlands) package for data visualization.^27^

## 3. RESULTS

### 3.1 Selection of IVs

After screening, there were 26 different types of gut microbiota and 6 different types of inflammatory proteins with potential causal relationships with PTD. The F-statistics for all IVs were above 10, indicating no evidence of weak instrument bias.^28^ After the Bonferroni adjustment, the p-values were all below the Bonferroni threshold.

### 3.2 Causal association between gut microbiota and PTD

When evaluating the causal association of gut microbiota with PTD, 26 types of gut microbiota show potential causal relationships. Relevant details can be found in Supplementary Table S2. Among them, the abundances in stool of Thioalkalivibrionaceae (OR=0.816; 95%CI=0.690-0.966; p=0.018), Syntrophorhabdaceae (OR=0.832; 95%CI=0.714-0.970; p=0.018), Dorea (OR=0.883; 95%CI=0.789-0.988; p=0.030), Johnsonella ignava (OR=0.884; 95%CI=0.793-0.986; p=0.027), Butyricimonas sp900258545 (OR=0.924; 95%CI=0.874-0.977; p=0.005), Turicibacter (OR=0.936; 95%CI=0.887-0.987; p=0.014), Turicibacteraceae (OR=0.950; 95%CI=0.903-0.999; p=0.045), and CAG-882 sp003486385 (OR=0.952; 95%CI=0.918-0.987; p=0.008) show a negative association with PTD. This indicates that an increase in the abundance of these microbes leads to a decreased risk of PTD.

Conversely, the following types of gut microbiota show a positive association with PTD: Bifidobacterium bifidum (OR=1.049; 95%CI=1.013-1.087; p=0.008), CAG-448 (OR=1.051; 95%CI=1.005-1.099; p=0.031), Succinivibrio (OR=1.059; 95%CI=1.011-1.110; p=0.016), Coprobacillus (OR=1.065; 95%CI=1.009-1.125; p=0.022), Succinivibrionaceae (OR=1.066; 95%CI=1.004-1.132; p=0.036), Lawsonibacter sp900066645 (OR=1.082; 95%CI=1.004-1.165; p=0.039), CAG-698 (OR=1.092; 95%CI=1.010-1.181; p=0.027), Brachyspira (OR=1.094; 95%CI=1.008-1.188; p=0.032), Treponema D (OR=1.095; 95%CI=1.011-1.187; p=0.026), UBA11963 (OR=1.105; 95%CI=1.019-1.198; p=0.015), Elusimicrobiaceae (OR=1.106; 95%CI=1.012-1.209; p=0.027), Prevotella sp900318625 (OR=1.111; 95%CI=1.030-1.197; p=0.006), UBA7748 sp900314535 (OR=1.120; 95%CI=1.016-1.233; p=0.022), Fournierella massiliensis (OR=1.121; 95%CI=1.010-1.245; p=0.032), Blautia A sp002159835 (OR=1.123; 95%CI=1.035-1.219; p=0.005), CAG-465 sp000433135 (OR=1.125; 95%CI=1.042-1.215; p=0.003), An7 (OR=1.139; 95%CI=1.028-1.262; p=0.013), and Thermoplasmatota (OR=1.190; 95%CI=1.038-1.364; p=0.012).

When evaluating the causal effects of PTD on the gut microbiota, it was observed that all p-values were greater than 0.05, suggesting that PTD does not have an effect on the gut microbiota under consideration. The relevant details are presented in Supplementary Table S3. The final analysis reveals potential causal relationships between 26 types of gut microbiota and PTD, as illustrated in Figure 2.

**Figure 2.**
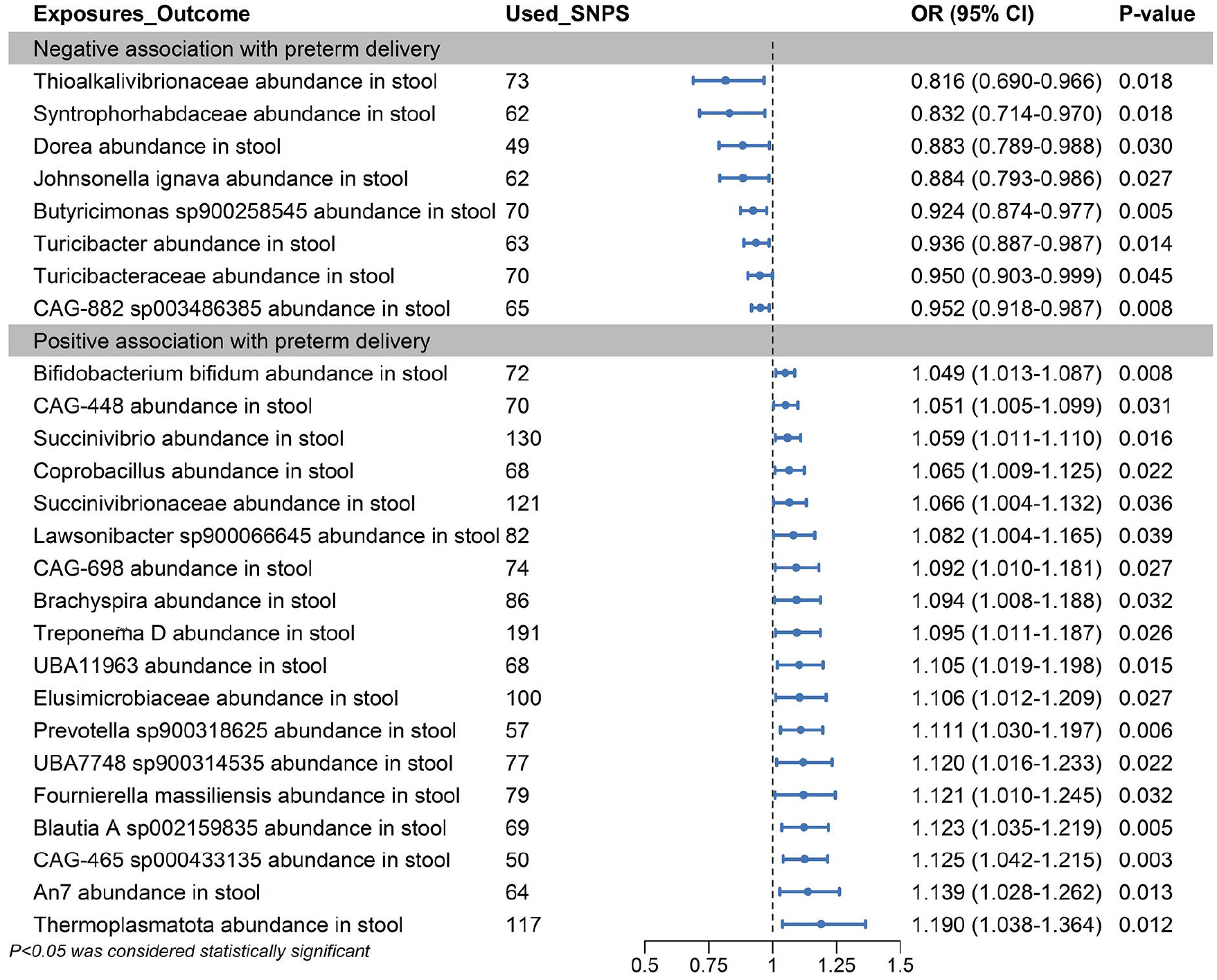
Mendelian randomization analyses show causal effects between gut microbiota and preterm delivery. The forest plot shows significant causal associations with p<0.05 and the estimated odds ratio (OR) with 95% confidence intervals(CI).

### 3.3 Causal association between inflammatory proteins and PTD

When evaluating the causal association of inflammatory proteins with PTD, 6 types of inflammatory proteins show potential causal relationships. Relevant details are provided in Supplementary Table S4. Among them, three inflammatory proteins exhibit a negative association with PTD: Interleukin-20 receptor subunit alpha (OR=0.842; 95%CI=0.738-0.960; p=0.010), C-C motif chemokine 19 (OR=0.907; 95%CI=0.847-0.971; p=0.005), and C-C motif chemokine 23 (OR=0.945; 95%CI=0.893-0.999; p=0.047). This indicates that an increase in these inflammatory proteins is associated with a decreased risk of PTD, suggesting that these inflammatory proteins may be protective factors against PTD.

The other three types of inflammatory proteins show a positive correlation with PTD. The IVW analysis results for these proteins are as follows: Interleukin-15 receptor subunit alpha (OR=1.058; 95%CI=1.000-1.119; p=0.050), C-X-C motif chemokine 10 (OR=1.100; 95%CI=1.008-1.199; p=0.032), and Interleukin-17A (OR=1.118; 95%CI=1.001-1.249; p=0.047) indicating that these inflammatory factors are risk factors for PTD.

When evaluating the causal effects of PTD on the inflammatory proteins, it was observed that all p-values were greater than 0.05, suggesting that PTD does not have an effect on the inflammatory proteins under consideration. The relevant details are provided in Supplementary Table S5. The final analysis reveals potential causal relationships between 6 types of inflammatory proteins and PTD, as illustrated in Figure 3.

**Figure 3.**
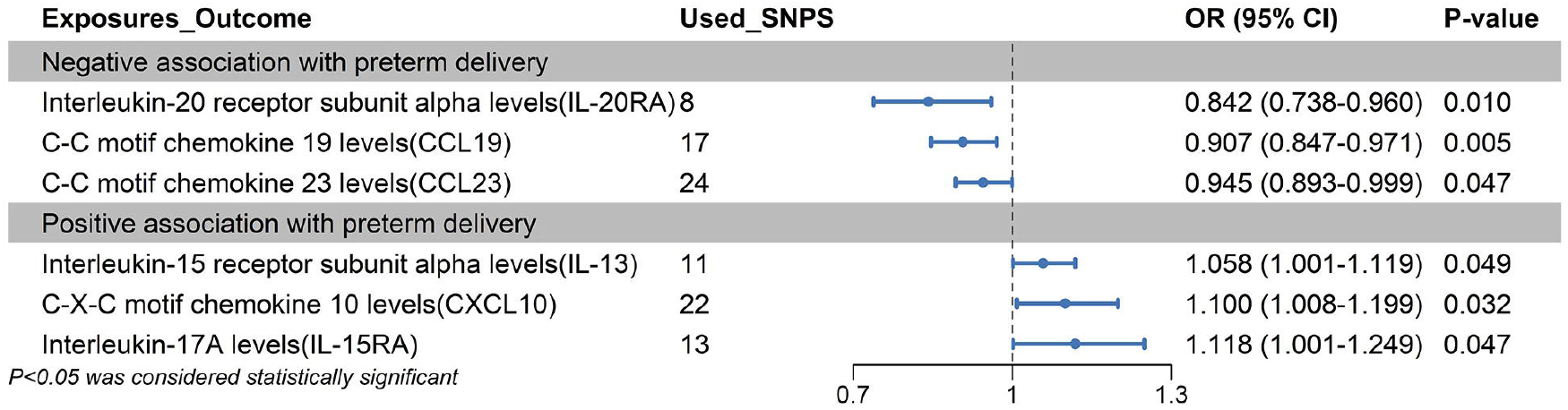
Mendelian randomization analyses show causal effects between inflammatory proteins and preterm delivery. The forest plot shows significant causal associations with p<0.05 and the estimated odds ratio (OR) with 95% confidence intervals (CI).

### 3.4 Mediation MR analyses of inflammatory proteins

In the 2SMR analysis, 26 types of gut microbiota and 6 types of inflammatory proteins were found to be causally associated with PTD, of which the abundance of Elusimicrobiaceae in stool was significantly associated with the C-C motif chemokine 23 levels (CCL23). Similarly, the abundance of Thioalkalivibrionaceae in stool was significantly associated with the Interleukin-20 receptor subunit alpha levels (IL-20RA). The results are presented in Supplementary Table S6. Genetically predicted Elusimicrobiaceae abundance in stool was negatively associated with the CCL23 (IVW; β=-0.090, SE=0.042, P=0.034). Genetically predicted Thioalkalivibrionaceae abundance in stool was positively associated with the IL-20RA levels (MR Egger; β=0.411, SE=0.188, P=0.032, IVW; β=0.199, SE=0.088, P=0.023).

When evaluating the causal effects of inflammatory proteins on gut microbiota, it was observed that all p-values were greater than 0.05, suggesting that inflammatory proteins do not affect the gut microbiota under consideration. The relevant details are provided in Supplementary Table S7.

We found that the abundance of Elusimicrobiaceae in stool was associated with decreased levels of CCL23, and this decrease was associated with an increased risk of PTD. Our study showed that CCL23 accounted for 5.09% (95%CI; 4.1%-8.7%) of the increased risk associated with the abundance of Elusimicrobiaceae in stool concerning the CCL23 and PTD. These results preliminarily illustrate that Elusimicrobiaceae abundance in stool may increase the risk of PTD partially by diminishing the effects of the CCL23 on PTD.

Similarly, we found that the abundance of Thioalkalivibrionaceae in stool was associated with increased levels of IL-20RA, and this increase was associated with a decreased risk of PTD. Our study showed that the IL-20RA accounted for 16.88% (95%CI; 12.77%-20.99%) of the increased risk associated with the abundance of Thioalkalivibrionaceae in stool concerning the IL-20RA and PTD. These results preliminarily illustrate that Thioalkalivibrionaceae abundance in stool may decrease the risk of PTD partially by enhancing the effects of the IL-20RA on PTD. The results are presented in Supplementary Table S8, and the schematic representation is shown in Figure 4.

**Figure 4.**
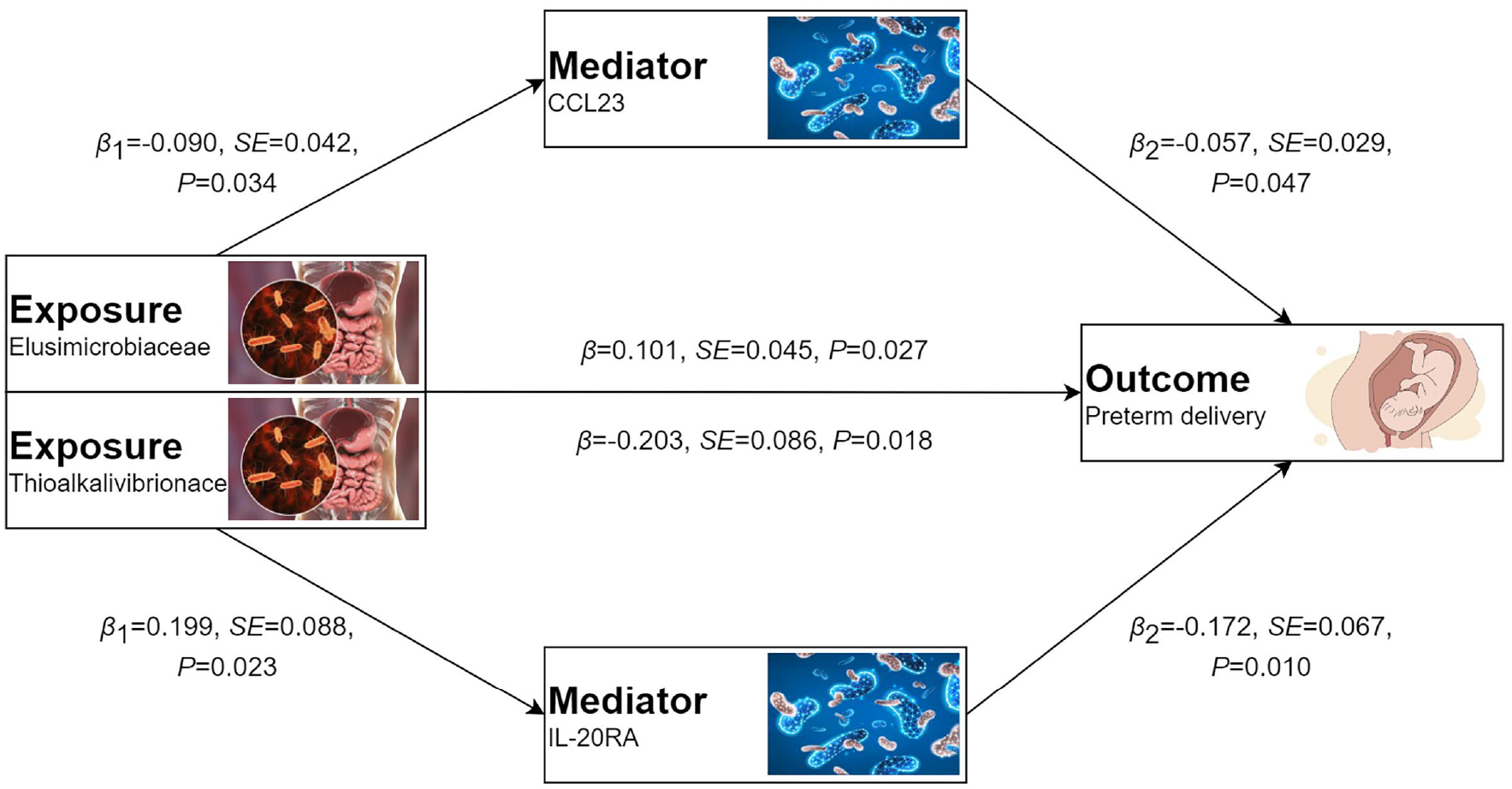
Schematic diagram of the mediating effect of Inflammatory Proteins between Gut microbiota and Preterm delivery.

### 3.5 Sensitivity analysis

To address potential pleiotropy in our causal effect estimates, we performed multiple sensitivity analyses. Cochran’s Q-test and funnel plot analysis revealed no evidence of heterogeneity or asymmetry among the SNPs involved in the causal relationship. Furthermore, leave-one-out analysis validated the impact of each SNP on the overall causal estimates. Moreover, re-analysis of the MR study after excluding individual SNPs consistently yielded similar results, indicating that all SNPs contributed significantly to the establishment of the causal relationship, with relevant details provided in Supplementary Table S9.

## 4. Discussion

In the present study, we investigated the causal association among 473 gut microbiota phenotypes, 91 inflammatory proteins, and PTD using large-scale genetic data and MR analysis. To our knowledge, this is the first MR analysis to explore the causal relationships among various gut microbiota phenotypes, inflammatory proteins, and PTD. Through stringent inclusion criteria and sensitivity analyses, we have identified potential causal links between 26 distinct gut microbiota types and PTD, with 2 specific inflammatory proteins potentially serving as mediators in this pathway.

Our study highlights significant associations between various types of gut microbiota and PTD, contributing to the body of evidence regarding the role of the gut microbiome in PTD. We identified 26 gut microbiota species with potential causal relationships to PTD, which opens avenues for further exploration. Several taxa exhibited negative associations with PTD. Specifically, higher abundances in the stool of Thioalkalivibrionaceae, Syntrophorhabdaceae, Dorea, Johnsonella ignava, Butyricimonas sp900258545, Turicibacter, Turicibacteraceae, and CAG-882 sp003486385 were linked to a reduced risk of PTD. This suggests that higher abundances of these microbes may confer a protective effect against PTD. The observed odds ratios (OR) for these communities (e.g., Thioalkalivibrionaceae OR=0.816) indicate a statistically significant reduced risk of PTD, highlighting the potential role of gut microbiota in modulating pregnancy outcomes. This observation aligns with a growing literature that suggests healthy gut microbiota may positively influence pregnancy by enhancing immune function and reducing inflammation.^29-30^ Conversely, certain taxa demonstrated positive associations with PTD, including Bifidobacterium bifidum, CAG-448, Succinivibrio, Coprobacillus, Succinivibrionaceae, Lawsonibacter sp900066645, CAG-698, Brachyspira, Treponema D, UBA11963, Elusimicrobiaceae, Prevotella sp900318625, UBA7748 sp900314535, Fournierella massiliensis, Blautia A sp002159835, CAG-465 sp000433135, An7, and Thermoplasmatota. This Underscore the potential of these taxa to contribute to increased PTD risk. It is possible that dysbiosis, characterized by an abundance of pro-inflammatory or pathogenic microbes, may disrupt normal physiological processes during pregnancy, thereby raising the likelihood of PTD.^31-32^ The mechanisms underlying these associations warrant further investigation, including exploring how these microbial communities could influence metabolic pathways, maternal immune responses, and the overall uterine environment. Moreover, our results indicate that PTD itself does not appear to have a significant impact on the above gut microbiota, as evidenced by non-significant p-values across the evaluated microbiota types. This suggests a unidirectional relationship where gut microbiota potentially influences the course of pregnancy rather than the reverse. This finding emphasizes the necessity to consider gut health in prenatal care strategies and interventions aimed at preventing PTD.

The findings of this study shed light on the complex interplay between inflammatory proteins and PTD, highlighting the potential of specific inflammatory markers as predictive biomarkers and therapeutic targets for PTD. Our analysis identified 6 distinct types of inflammatory proteins associated with PTD, with several demonstrating noteworthy causal relationships. Notably, a subset of these proteins, including IL-20RA, C-C motif chemokine 19, and CCL23, exhibited a negative association with PTD. The observed OR for these proteins (e.g., IL-20RA OR=0.842) indicate a statistically significant reduced risk of PTD, suggesting that higher levels of these inflammatory factors may confer a protective effect. These findings align with emerging evidence that suggests certain inflammatory pathways may play a beneficial role in pregnancy by modulating immune responses and maintaining a healthy intrauterine environment. For instance, IL-20RA has been linked to anti-inflammatory and tissue-repairing functions, which could contribute to the observed protective effect against PTD.^33^ Similarly, the chemokines C-C motif chemokine 19 and CCL23 have been implicated in the recruitment and regulation of immune cells, potentially promoting a balanced inflammatory state during pregnancy. ^34^ In contrast, the remaining three inflammatory proteins, including Interleukin-15 receptor subunit alpha, C-X-C motif chemokine 10, and Interleukin-17A, exhibited a positive association with PTD. The elevated odds ratios (e.g., Interleukin-17A OR=1.118) for these factors suggest that they may contribute to an increased risk of PTD. These pro-inflammatory mediators have been linked to various pathological processes, such as uterine inflammation, placental dysfunction, and premature cervical ripening, which could potentially disrupt the normal course of pregnancy and lead to PTD.^35^ Our study similarly found that PTD itself does not have a significant impact on the inflammatory proteins under investigation, as evidenced by non-significant p-values. This finding suggests a unidirectional relationship, where specific inflammatory profiles may precede and potentially influence the risk of PTD rather than the reverse. This observation underscores the importance of considering inflammatory biomarkers in the context of prenatal care and the development of targeted interventions.

The comprehensive analysis of our study has uncovered intricate relationships between gut microbiota, inflammatory proteins, and PTD, providing valuable insights into the complex interplay of these factors. The identification of specific microbial taxa and inflammatory proteins with causal associations to PTD risk is a significant step forward in understanding the underlying mechanisms driving this adverse pregnancy outcome. Notably, the abundance of Elusimicrobiaceae and Thioalkalivibrionaceae in stool exhibited significant associations with specific inflammatory proteins, namely CCL23 and IL-20RA, respectively. The negative association between genetically predicted Elusimicrobiaceae abundance and CCL23 levels suggests that higher levels of this microbiota may correlate with decreased levels of this pro-inflammatory chemokine. Our analysis indicated that this decrease was associated with an increased risk of PTD, with CCL23 accounting for 5.09% of the increased risk linked to the abundance of Elusimicrobiaceae. This finding reveals a potential mechanism by which diminished levels of CCL23 may contribute to PTD risk, suggesting that Elusimicrobiaceae abundance in stool may impair the protective effects of this inflammatory marker. Conversely, the analysis revealed that the abundance of Thioalkalivibrionaceae in stool was positively correlated with increased levels of IL-20RA, which in turn was associated with a decreased risk of PTD. This relationship was elucidated by our finding that IL-20RA accounted for 16.88% of the increased risk associated with Thioalkalivibrionaceae abundance concerning PTD. This illustrates that higher levels of Thioalkalivibrionaceae may mitigate the risk of PTD by enhancing the protective effects of IL-20RA. These results suggest foundational patterns in the interactions between gut microbiota and inflammatory markers in the context of pregnancy and PTD. This reinforces the hypothesis that modulation of gut microbiota can influence systemic inflammatory responses and thereby impact pregnancy outcomes.^36^ In addition, when we evaluated the causal effects of these two inflammatory proteins on gut microbiota, all p-values were greater than 0.05, indicating that the inflammatory proteins we studied do not significantly affect the composition of gut microbiota. This unidirectional relationship suggests that gut microbiota may play a more pronounced role in affecting inflammatory profiles rather than being influenced by them, which has implications for understanding the etiology of PTD.

The present study has several strengths that can guide future directions in PTD research. First, our work comprises the first systematic investigation to examine the causal relationship between gut microbiota, inflammatory proteins, and PTD. This unique contribution is underscored by the comprehensive analysis of 473 distinct types of gut microbiota and 91 inflammatory proteins. By using an MR design, the study effectively mitigated issues related to reverse causality and residual confounding variables. Furthermore, thorough sensitivity analyses were conducted to eliminate the potential influence of genetic polymorphisms, thereby enhancing the validity of the causal inferences drawn from the study. Second, we have identified a set of gut microbiota and inflammatory proteins that exhibit a strong correlation with the risk of PTD. These findings present potential biomarkers for non-invasive stool testing in PTD that warrant validation through subsequent experiments. Finally, we have proposed a potential axis of Elusimicrobiaceae–CCL23–PTD, as well as another axis Thioalkalivibrionaceae–IL-20RA–PTD, which may serve as a foundation for drug development strategies in PTD research. Further investigation utilizing in vitro and in vivo models is necessary to validate these proposed axes and to develop targeted therapies accordingly. which may serve as a foundation for drug development strategies in PTD research.

This study conducted TSMR, BDMR, and 2SMR analysis utilizing large GWAS datasets, demonstrating high statistical efficiency. The conclusions drawn in our study are grounded in genetic IVs, with causal inference conducted through multiple MR analysis methods. The findings are robust and unaffected by horizontal pleiotropy and other confounding factors. However, it is important to acknowledge several limitations in our study. Firstly, our analysis was performed using the European population, limiting its application to everyone worldwide.^37^ Second, the dataset on PTD is general and lacks subtype information, which may not accurately reflect the characteristics of specific subtypes within PTD. Third, using summary-level statistics rather than individual-level data in our analysis limits our ability to investigate causal relationships within specific subgroups, such as females and males. Moreover, despite efforts to detect and remove outlier variants, the potential influence of horizontal pleiotropy on our findings cannot be completely ruled out.^38^ Finally, our research indicates a modest genetic prediction of PTD, mediated by the CCL23 at 5.09% and the IL-20RA at 16.88%, suggesting the need for further investigation into additional mediators.

## 5. Conclusions

This study demonstrates a causal relationship between specific gut microbiota and inflammatory proteins and PTD through MR analyses. Notably, Elusimicrobiaceae and Thioalkalivibrionaceae were significantly associated with PTD, with mediation occurring via CCL23 and IL-20RA, respectively. The findings suggest that targeting these microbiota and inflammatory pathways could offer potential avenues for intervention in preventing PTD. However, further research is needed to explore the underlying mechanisms and validate these associations in clinical settings.

## Data Availability

The analysis utilized publicly available datasets. Detailed information on all original contributions can be found in the "Data Sources" section, including specific download links and accession numbers. Readers can refer to this section for access. For further inquiries, please contact the corresponding author.

https://www.ebi.ac.uk/gwas/

http://egg-consortium.org/

## 6. ETHICAL APPROVAL

This research utilized publicly accessible data from GWAS studies. Each individual study included in the GWAS was approved by the respective Institutional Review Board, and participants or their authorized representatives provided informed consent.

## 7. AUTHORS CONTRIBUTIONS

Min Zhang conducted the data analysis and interpretation and wrote the manuscript. Xiaodan Chen and Yan Zhang were responsible for proposing the research hypothesis. Jia Huang and Ling Chen were responsible for conceptualizing the study.

